# Regional Difference in Seroprevalence of SARS-CoV-2 in Tokyo: Results from the community point-of-care antibody testing

**DOI:** 10.1101/2020.06.03.20121020

**Authors:** Morihito Takita, Tomoko Matsumura, Kana Yamamoto, Erika Yamashita, Kazutaka Hosoda, Tamae Hamaki, Eiji Kusumi

## Abstract

The serosurvey is an alternative way to know the magnitude of the population infected by coronavirus disease 2019 (COVID-19) since the expansion of capacity of the polymerase chain reaction (PCR) to detect the severe acute respiratory syndrome coronavirus 2 (SARS-CoV-2) was delayed. We herein report seroprevalence of COVID-19 accessed in the two community clinics in Tokyo. The point-of-care immunodiagnostic test was implemented to detect the SARS-CoV-2 specific IgG antibody in the peripheral capillary blood. The overall positive percentage of SARS-CoV-2 IgG antibody is 3.83% (95% confidence interval: 2.76-5.16) for the entire cohort (*n* =1,071). The central Tokyo of 23 special wards exhibited a significantly higher prevalence compared to the other area of Tokyo (*p* =0.02, 4.68% [95%CI: 3.08-6.79] versus 1.83 [0.68-3.95] in central and suburban Tokyo, respectively). The seroprevalence of the cohort surveyed in this study is low for herd immunity, which suggests the need for robust disease control and prevention. A community-based approach, rather than state or prefectural levels, is of importance to figure out profiles of the SARS-COV-2 outbreak.

## Introduction

A current challenge for the measurement of the future outbreak of coronavirus disease 2019 (COVID-19) is to find a more focused or personalized strategy of infection control such as contact tracing until the establishment of the herd immunity by the previous infection or vaccination which is under development.^1^ The New York State announced that the seroprevalence in lower-income communities was higher than the overall population (27 versus 20% of positive for IgG of the severe acute respiratory syndrome coronavirus 2, SARS-CoV-2, respectively).^2^ We already described our interim analysis of point-of-care antibody testing for COVID-19, conducted in the community clinics in Tokyo, Japan.^3^ We herein reported our extensive analysis which showed unique geographical characteristics of seroprevalence of COVID-19 in Tokyo.

## Methods

The Institutional Review Board of Navitas Clinic approved the present study (Approval Number: NC2020-01). Asymptomatic subjects have been recruited by web posting of our clinic, and written consent was obtained from all participants prior to the test. The present study here is an observational study analyzing data collected from the medical record. The study participants paid the cost of the point-of-care test since no insurance and public funding was not available to defray it in Japan.

The SARS-CoV-2 specific IgG antibody in the peripheral capillary blood was detected by the point-of-care immunodiagnostic test (SARS-CoV-2 Antibody Testing Kit IgG RF-NC002, Kurabo Industries Ltd, Osaka, Japan) in the two community clinics located in the major railway stations in Tokyo (Navitas Clinic Shinjuku and Tachikawa). The rapid test implemented in this study indicated 76.4, 100 and 94.2% of positive, negative and overall agreement rates, respectively, in accordance with the manufacturer’s investigation.

The 95% exact binomial confidential interval (CI) was calculated with the method of Clopper-Pearson using PASS 14 (NCSS, LLC. Kaysville, UT). Statistical significance was considered when two-side *p*-value <0.05. The descriptive and Fisher’s exact test was performed with IBM SPSS Statistics 26 (IBM Corp., Armonk, NY)

## Results

The overall positive percentage of SARS-CoV-2 IgG antibody is 3.83% (95% CI: 2.76-5.16) for the entire cohort (*n* =1,071) (Table 1). No trend of increase in the positive proportion by time was observed (Supplemental Table 1). All participants who had a history of SARS-CoV-2 PCR positive (*n* =5) showed positive results of IgG, whereas IgG positive was also seen in two with the PCR negative. The seroprevalence in central Tokyo of 23 special wards was significantly higher than the other area of Tokyo after classification by residence *(p* =0.02, 4.68% [95%CI: 3.08-6.79] versus 1.83 [0.68-3.95] in central and suburban Tokyo, respectively). Furthermore, we found the regional trend inside the central Tokyo; the highest and lowest prevalence was found in the Southern and Northern areas (7.92% [95%CI: 3.48-15.01] and 0 [0.00-5.13]), respectively, although no statistical significance was observed. (Supplementary Table 2 and Supplementary Figure for classification of regions).

**Table 1.** Characteristics of Study Participants and Proportion with SARS-CoV-2 IgG Positive. Neighboring prefectures of Tokyo include Kanagawa, Saitama, Chiba and Yamanashi Prefectures.

| Characteristics | Sample size | Proportion of sample, % (95%CI) | No. positive | Proportion positive for SARS-CoV-2 IgG, % (95%CI) |
| --- | --- | --- | --- | --- |
| Entire sample | 1,071 | 100 | 41 | 3.83 (2.76-5.16) |
| Sex |  |  |  |  |
| Male | 576 | 53.78 (50.74-56.80) | 22 | 3.82 (2.41-5.73) |
| Female | 495 | 46.22 (43.20-49.26) | 19 | 3.84 (2.33-5.93) |
| Age, year |  |  |  |  |
| ≤17 | 13 | 1.21 (0.65-2.07) | 0 | 0 |
| 18-34 | 134 | 12.51 (10.59-14.64) | 11 | 8.21 (4.17-14.21) |
| 35-54 | 653 | 60.97 (57.98-63.91) | 19 | 2.91 (1.76-4.51) |
| ≥55 | 271 | 25.30 (22.72-28.02) | 11 | 4.06 (2.04-7.15) |
| Episodes of fever after Dec 2019 | 332 | 31.00 (28.24-33.87) | 19 | 5.72 (3.48-8.79) |
| History of PCR test for SARS-CoV-2 | 45 | 4.20 (3.08-5.58) | 7 | 15.56 (6.49-29.46) |
| PCR *positive* | 5 | 0.47 (0.15-1.09) | 5 | 100 |
| PCR *negative* | 40 | 3.73 (2.68-5.05) | 2 | 5.00 (0.61-16.92) |
| Cohabitant diagnosed with COVID-19 | 9 | 0.84 (0.38-1.59) | 4 | 44.44 (13.70-78.80) |
| Healthcare worker | 175 | 16.34 (14.17-18.69) | 7 | 4.00 (1.62-8.07) |
| Residence |  |  |  |  |
| Central Tokyo; 23 special wards | 565 | 52.75 (49.71-55.78) | 27 | 4.78 (3.17-6.88) |
| Suburban Tokyo | 317 | 29.60 (26.88-32.43) | 5 | 1.58 (0.51-3.64) |
| Neighboring prefectures of Tokyo | 173 | 16.15 (14.00-18.50) | 9 | 5.20 (2.41-9.65) |
| Other | 16 | 1.49 (0.86-2.41) | 0 | 0 |

## Discussion

The overall seroprevalence in this study is low as the survey in Los Angeles,^4^ which suggests that the majority of the population is immunologically naïve for SARS-CoV-2. Naturally, the prevalence is higher in the place with a higher density of population, such as central Tokyo, since the SARS-CoV-2 is primarily transmitted by droplets. The regional trend of seroprevalence is similar to the cumulative number of COVID-19 patients per unit population (Supplementary Table 3). These facts suggest that the community-based investigation would be beneficial to explore the cause of epidemic contagion. Limitation includes the selection bias and accuracy of the test kit. Concerns about the risk of COVID-19 infection such as past fever, illness of cohabitants or co-workers and working environment were common reasons for the participation of this study, which can cause elevation of seroprevalence. Less sensitivity of test kit develops the underestimation of the prevalence. Further serosurvey, along with the fine characterization of population, is warranted for the measurement of a future outbreak.

## Data Availability

The data that support the findings of this study are available from the corresponding author upon reasonable request to protect the privacy of study participants.

## Additional Contributions

We thank the participants and clinical collaborators involved in performing point-of-care tests.

## External Funding Source

None. The present study was performed by intramural funding of Navitas Clinic.

